# Recovery from Covid-19 critical illness: a secondary analysis of the ISARIC4C CCP-UK cohort study and the RECOVER trial

**DOI:** 10.1101/2021.06.15.21258879

**Authors:** Ellen Pauley, Thomas M Drake, David M Griffith, Nazir I Lone, Ewen M Harrison, J Kenneth Baillie, Janet T Scott, Timothy S Walsh, Malcolm G Semple, Annemarie B Docherty, on behalf of the ISARIC4C and RECOVER Investigators

## Abstract

**Background:** We aimed to compare the prevalence and severity of fatigue in survivors of Covid-19 versus non-Covid-19 critical illness, and to explore potential associations between baseline characteristics and worse recovery.

**Methods:** We conducted a secondary analysis of two prospectively collected datasets. The population included was 92 patients who received invasive mechanical ventilation (IMV) with Covid-19, and 240 patients who received IMV with non-Covid-19 illness before the pandemic. Follow-up data was collected post-hospital discharge using self-reported questionnaires. The main outcome measures were self-reported fatigue severity and the prevalence of severe fatigue (severity >7/10) 3 and 12-months post-hospital discharge.

**Results:** Covid-19 IMV-patients were significantly younger with less prior comorbidity, and more males, than pre-pandemic IMV-patients. At 3-months, the prevalence (38.9% [7/18] vs. 27.1% [51/188]) and severity (median 5.5/10 vs. 5.0/10) of fatigue was similar between the Covid-19 and pre-pandemic populations respectively. At 6-months, the prevalence (10.3% [3/29] vs. 32.5% [54/166]) and severity (median 2.0/10 vs. 5.7/10) of fatigue was less in the Covid-19 cohort. In the Covid-19 population, women under 50 experienced more severe fatigue, breathlessness, and worse overall health state compared to other Covid-19 IMV-patients. There were no significant sex differences in long-term outcomes in the pre-pandemic population. In the total sample of IMV-patients included (i.e. all Covid-19 and pre-pandemic patients), having Covid-19 was significantly associated with less severe fatigue (severity <7/10) after adjusting for age, sex, and prior comorbidity (adjusted OR 0.35 (95%CI 0.15-0.76, p=0.01).

**Conclusion:** Fatigue may be less severe after Covid-19 than after other critical illness.

## Introduction

Post Intensive Care Syndrome (PICS) describes the constellation of physical, psychological and cognitive symptoms affecting 25% of critical illness survivors, which include fatigue, muscle weakness, and posttraumatic stress^1^. PICS may persist for longer than 5 years and is associated with high hospital resource use and readmission rates^2^. Implementation of post-ICU rehabilitation guidelines is inconsistent, and the best ways to support intensive care unit (ICU) survivors are uncertain^3–5^; Identifying those at greatest risk of PICS and providing evidence-based strategies to enhance their recovery remain priorities for critical care research^5^. The Covid-19 pandemic heralded a dramatic increase in ICU admissions, over half of whom survived to discharge, thus a vast new cohort of patients at risk of PICS has emerged^6,7^.

Many patients who survive acute-Covid-19 experience persistent symptoms beyond 4 weeks, known as ‘long-Covid’^8^. This includes a wide range of symptoms, such as breathlessness, fatigue, muscle pain, many of which overlap with PICS and other post-viral syndromes^1,9–16^. The most common symptom of long-Covid described following both community and hospital-managed acute-Covid-19 was fatigue: fatigue was reported in 97.7% of community-managed cases who reported symptoms lasting over 28 days, and in 83% and 98% of UK and China patients respectively greater than 3-months after hospital discharge following admission with acute-Covid-19^11,16,17^. Recent studies have shown requiring invasive mechanical ventilation (IMV) is associated with experiencing worse fatigue and other recovery, but have lacked appropriate controls to compare ‘long-Covid’ to recovery from all critical illness^9,11,12^.

The prevalence of fatigue in ICU-survivors ranges from 13.8% to 80.9%^18^. This has a profound negative impact on survivors’ quality of life (QOL), and is one of the most prevalent and debilitating challenges of ICU-survivorship^18,19^. Fatigue may lead to delayed return to employment and pre-critical illness physical fitness, difficulty in completing activities of daily living, social isolation, depression, and many other negative sequalae ^18–20^. It is unclear if recovery is different in survivors of Covid-19 critical illness compared to other critical illness, requiring distinct risk assessment tools or interventions to support their recovery.

We aimed to characterise the prevalence and severity of fatigue in Covid-19 ICU-survivors, compared to survivors of non-Covid-19 critical illness, and to explore potential associations between baseline characteristics and fatigue severity in these populations to identify potential groups at greater risk.

## Methods

### Study design and population

This was a secondary analysis of two prospectively collected datasets.

#### Covid-19 cohort: International Severe Acute Respiratory Infection Consortium Coronavirus Clinical Characterisation Protocol - United Kingdom (ISARIC-4C CCP-UK)

Patients aged 18 years and over, admitted to hospital between 17^th^ January and 5^th^ October 2020 with confirmed or highly suspected SARS-CoV-2 infection at 31 UK centres, who consented to be contacted for follow-up studies, who completed a follow-up questionnaire, and who were discharged at least 90 days ago at the time of data collection were eligible for inclusion. Patients who did not survive to follow-up were excluded. We restricted this secondary analysis to patients who received IMV.

#### Pre-pandemic cohort: Evaluation of a Rehabilitation Complex Intervention for Patients Following Intensive Care Discharge (RECOVER) trial

Patients aged 18 and over, discharged from ICU between 1^st^ December 2010 and 31^st^ January 2013 at 2 hospitals in Edinburgh, Scotland, who received a minimum of 48-hours of IMV^21^. Patients randomised to the intervention group received enhanced hospital and community-based physical rehabilitation, the control group received routine care^21^. Both groups were included in this analysis, as there were no significant differences at baseline or follow-up^21^.

Full details of the populations included in CCP-UK and RECOVER, and study information, have previously been published^11,21^.

### Clinical variables

Both datasets captured demographic information, including age, sex, pre-existing comorbidities, and severity of acute illness. Variables indicating acute illness severity differed: CCP-UK included ISARIC-4C Mortality and World Health Organisation Severity Scores^22,23^; RECOVER included the Acute Physiology and Chronic Health Evaluation-II (APACHE-II) score^24^, duration of IMV, duration of ICU-admission, total and Respiratory Sequential Organ Failure Assessment (SOFA) scores^25^. Thus, and given the different underlying disease processes, acute illness severity was not directly compared. RECOVER captured survival at 3, 6, and 12-months post-hospital discharge: patients alive at each timepoint were contacted for follow-up.

### Outcomes

Both studies used patient-completed questionnaires to collect outcome data^11,21^. CCP-UK completed follow-up assessment via postal questionnaires (or telephone where this was not possible), which patients completed once and returned^11^. RECOVER completed follow-up assessment face-to-face or by telephone at 3-months, and by post at 6 and 12-months post-hospital discharge^21^.

In this secondary analysis, the primary outcome was patient-reported fatigue severity. Secondary outcomes were: ‘severe fatigue’, breathlessness, and measures of health-related QOL.

In both studies, fatigue severity was measured using a 10-point Visual Analogue Scale (VAS), where zero is no fatigue and ten is worst possible fatigue. ‘Severe fatigue’ was defined as fatigue of at least 7/10 on the VAS. CCP-UK used the Medical Research Council (MRC) Dyspnoea Scale^26^ to measure breathlessness, RECOVER used a 10-point VAS. The MRC Scale is a 1-5 stage scale measuring perceived respiratory disability: 1 being no breathlessness, 5 being unable to undertake activities of daily living due to breathlessness. CCP-UK measured QOL using the EuroQol (EQ5D-5L) instrument: this covers five dimensions: mobility, self-care, usual activities, pain/discomfort and anxiety/depression^27^. The person indicates their health state for each dimension. Summary EQ5D-5L indices were measured on a scale of 0 to 1: 1 being perfect health and 0 being worst health imaginable (typical population mean 0.87). RECOVER measured QOL using the 12-Item Short Form Survey (SF-12), subdivided into Summary scores of Mental and Physical Component Scores (MCS, PCS) (range:0-100; typical population mean:50, with higher values representing better health)^28^.

### Statistical methods

Covid-19 IMV-patients were compared to the pre-pandemic IMV-patients at baseline, follow-up, and potential associations with outcomes within each group were compared.

Covid-19 and pre-pandemic IMV-patients were compared at baseline in terms of age, sex, prior comorbidity, deprivation. Fatigue severity and the prevalence of severe fatigue was compared. Fatigue in Covid-19 IMV-patients was stratified by time since hospital discharge that patients responded to the CCP-UK questionnaire: approximately 3 (90-120 days) and 6-months (150-210 days), in order to compare fatigue at equivalent timepoints. Fatigue severity (VAS scores) and ‘severe fatigue’ were compared as continuous and binary outcomes respectively.

Where direct comparison was not possible, i.e. where the same variables measured with the same tools were not available in the two datasets, outcome variables from both datasets were presented separately.

Categorical data were summarised as frequencies and percentages, continuous data as median and interquartile range (IQR). To test for differences across comparison groups in categorical data, we used Fisher’s exact test and for continuous data, using the Wilcoxon rank-sum test for two-sample testing and Kruskall-Wallis where there were more than 2 groups.

We created linear regression models to adjust for age, sex, presence of comorbidities, and acute illness severity, for the outcomes of fatigue severity, breathlessness, and QOL in the Covid-19 and the pre-pandemic IMV-patient groups. Final model selection was guided by minimisation of the Akaike information criterion (AIC). Variables were only included in the model if they were present during the initial hospital admission. All models were checked for first order interactions and any meaningful interactions were retained and incorporated as dummy variables. In the Covid-19 cohort, we adjusted for the effects of age by sex, as this was identified as a significant interaction. Effect estimates are presented as mean differences alongside 95% confidence intervals (95%-CI). Statistical analyses were performed using R version 3.6.3 (R Foundation for Statistical Computing, Vienna, AUT) with the tidyverse, finalfit, eq5d and Hmisc packages. Statistical significance was taken at the level P≤0.05. Associations with outcomes were compared between the groups, for example to determine if age differences existed in both the Covid-19 and pre-pandemic groups.

We created an ordinal logistic regression and a logistic regression model including all patients (Covid-19 and pre-pandemic group), to adjust for age, sex, presence of comorbidities, and Covid-19 status on fatigue: with ordinal regression we explored fatigue in categories: 0-2/10, 2-4/10, 4-6/10, 6-8/10, 8-10/10, and with logistic regression we explored more severe reported fatigue, taking scores less than or equal to 7/10 or not as a binary outcome. Pre-pandemic outcomes at 6 months were included.

## Results

### Patients included

335 Covid-19 patients were included in the CCP-UK study: 27.5% (92/335) received IMV (Figure-1). 240 patients were included in RECOVER (pre-pandemic patients). A total of 332 patients (92 Covid-19, 240 pre-pandemic patients) who received IMV were included in this secondary analysis (Figure-1). 29.2% [70/240] and 35.0% [84/240] of the pre-pandemic cohort were admitted with Cardiovascular and Respiratory diagnosis categories respectively (Table-1).

**Figure 1:**
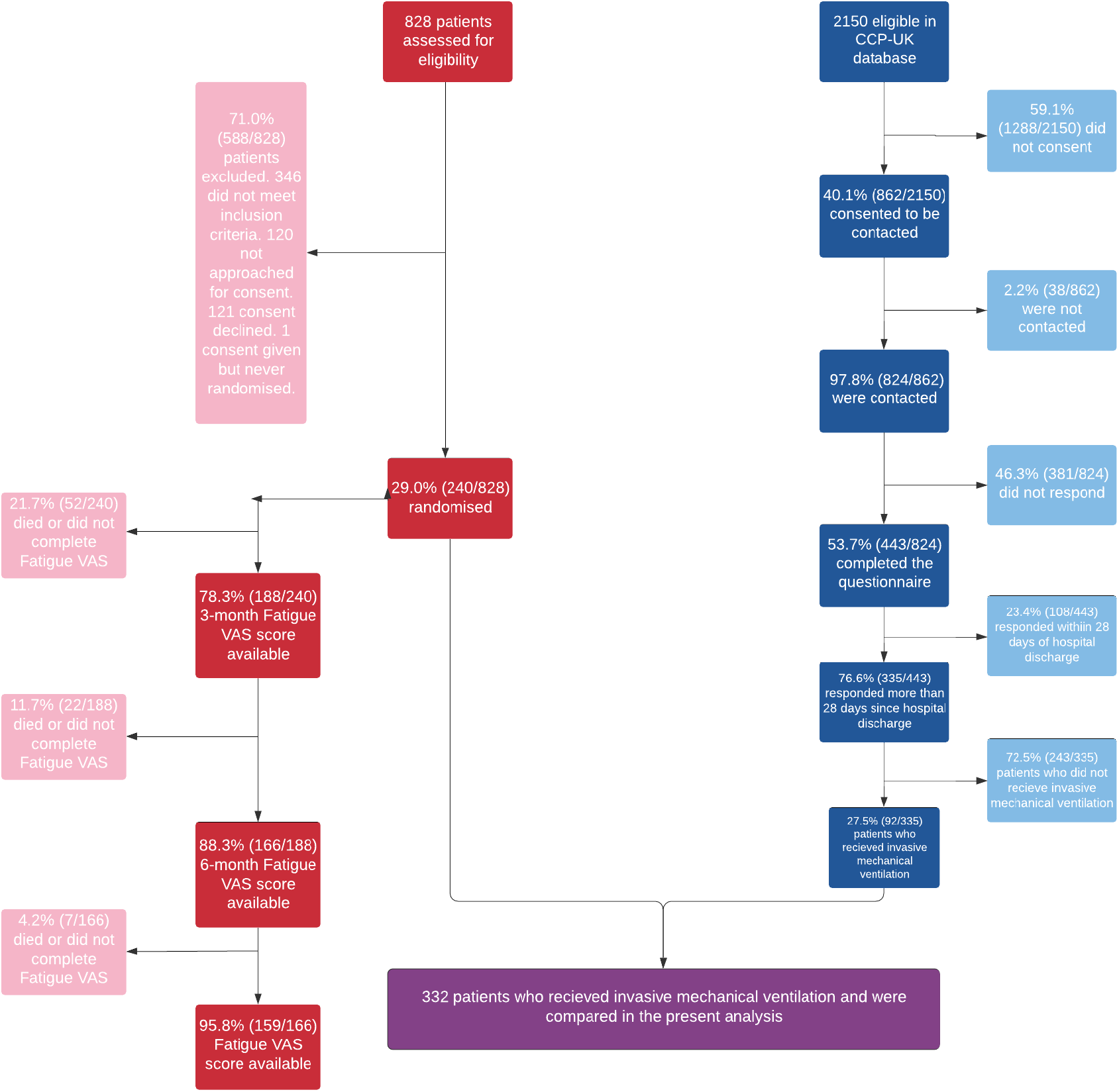
Patient inclusion flowchart. Red: RECOVER trial participants, n=240; Blue: ISARIC4C participants, n=92.

**Table 1:**
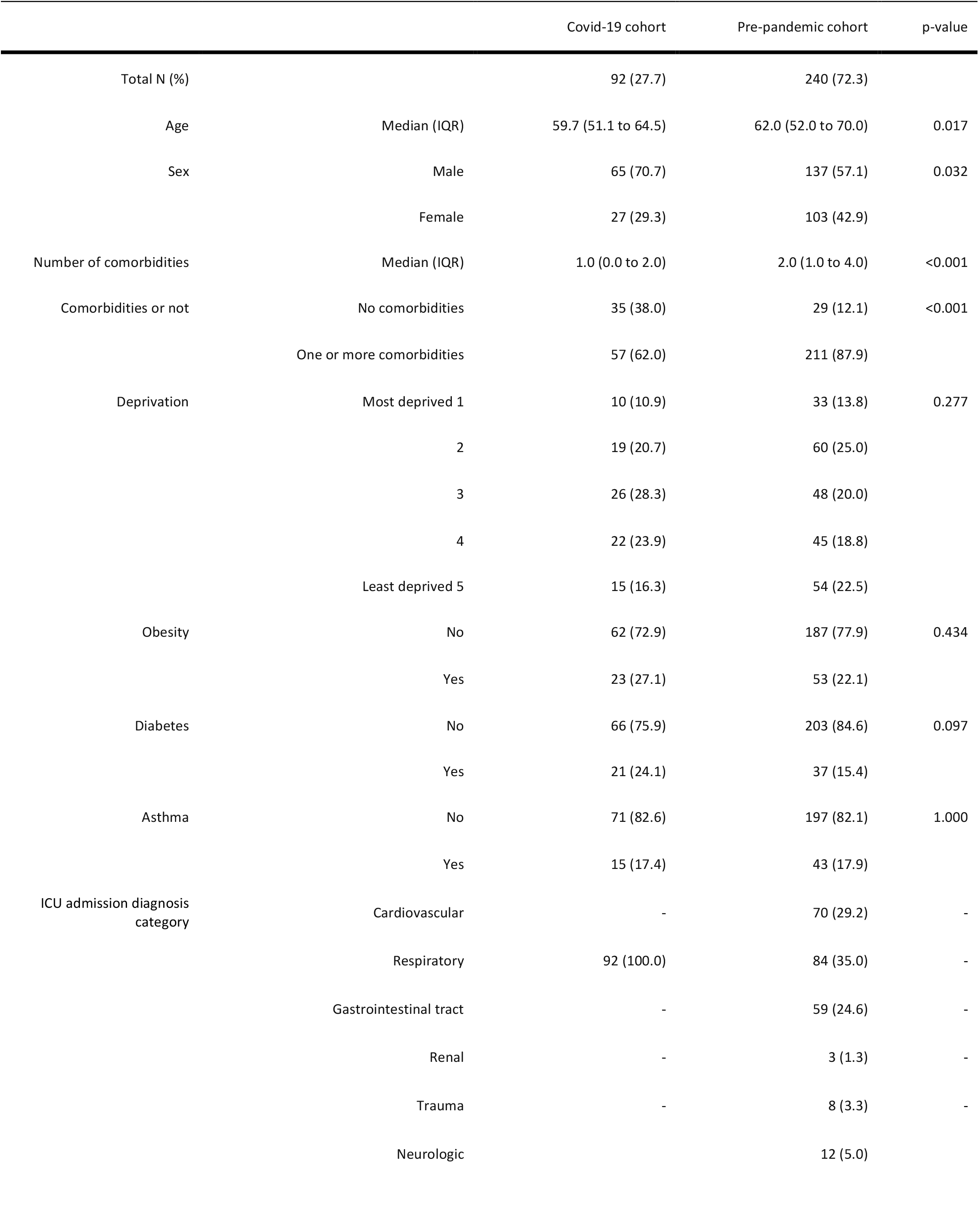
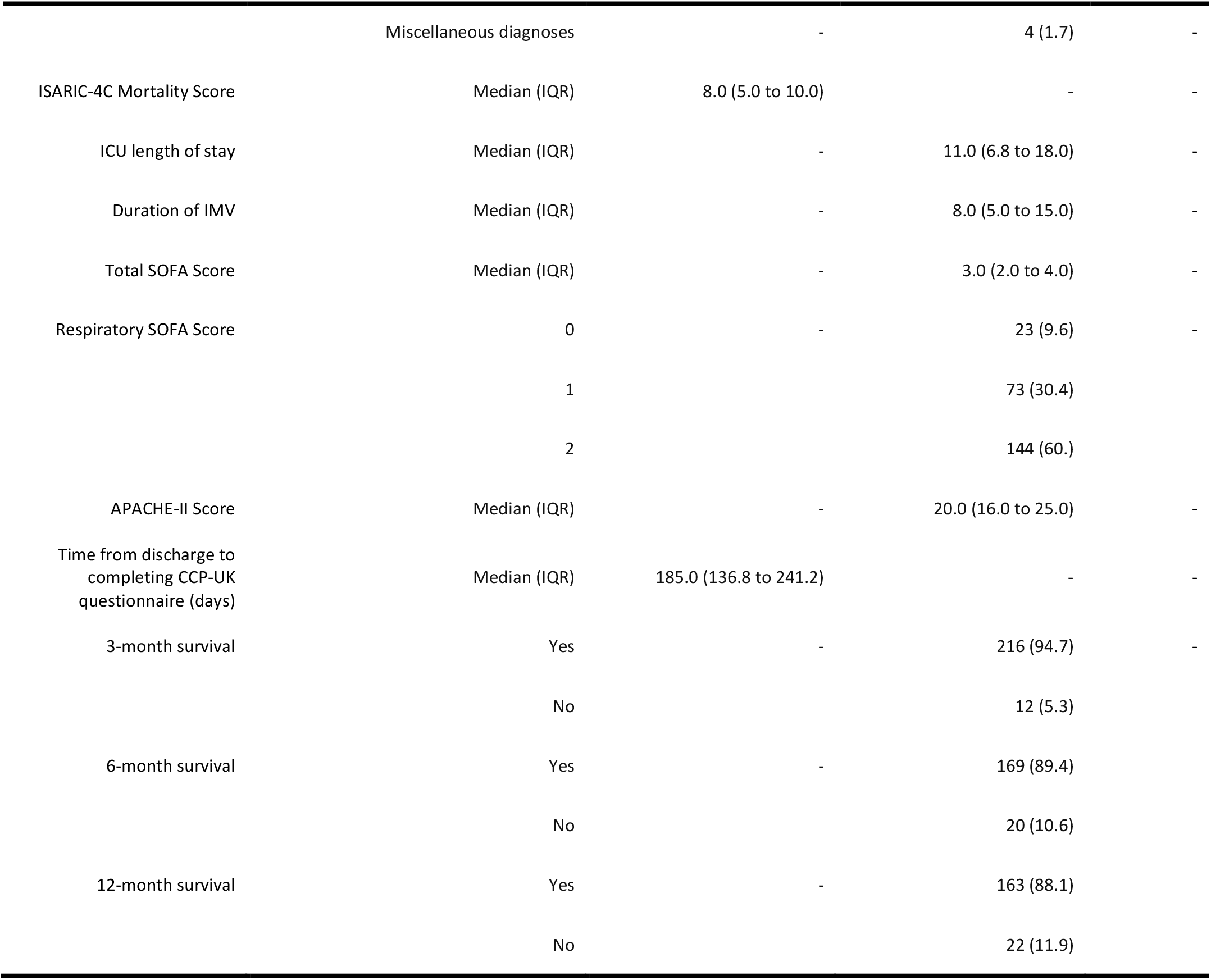
Patient characteristics of Covid-19 (n=92) and pre-pandemic IMV-patients (n=240). IQR – Interquartile range, presented as 25^th^ to 75^th^ centiles. Numbers are presented as N (%), unless otherwise denoted as a continuous variable. ICU – Intensive Care Unit. IMV – Invasive Mechanical Ventilation. SOFA – Sequential Organ Failure Assessment. APACHE-II – Acute Physiology and Chronic Health-II.

Covid-19 IMV-patients were significantly younger (median 59.7 years, IQR:51.1 to 64.5, p=0.017) than pre-pandemic IMV-patients (median 62.0 years, IQR:52.0 to 70.0, Table-1). A higher proportion of Covid-19 IMV-patients were males (70.7% [65/92]) than pre-pandemic (57.1% [137/240]). Significantly fewer Covid-19 IMV-patients had prior comorbidity (62.0% [57/92] vs 87.9% [211/240], p<0.001), and the median number of comorbidities was significantly lower (1, IQR:0 to 2 vs 2, IQR:1 to 4, p<0.001). Both groups were similar in terms of socioeconomic status, and the prevalence of obesity, diabetes, and asthma. Covid-19 IMV-patients responded a median 185 days, IQR:137 to 241, after hospital discharge (Table-1).

Pre-pandemic IMV-patients received a median 8 days of IMV, IQR:5 to 15; the median APACHE-II score was 20, IQR:16 to 25 (Table-1).

### Primary outcome

The median fatigue severity reported by Covid-19 IMV-patients at the timepoint they responded was 5.0, (n= 85), IQR:2.0 to 7.0, Table-2). At 3-months, fatigue severity was similar between Covid-19 (n=18) and pre-pandemic IMV-patients (n=188). At 6-months, pre-pandemic IMV-patients (n=166) reported significantly greater fatigue than Covid-19 IMV-patients (n=29), median 5.7/10, IQR:3.5 to 7.3/10 vs median 2/10, IQR:1.0 to 5.0/10, p<0.001). Fatigue at 12-months was not compared because Covid-19 IMV-patients had not accrued follow-up to this time point.

**Table 2:**
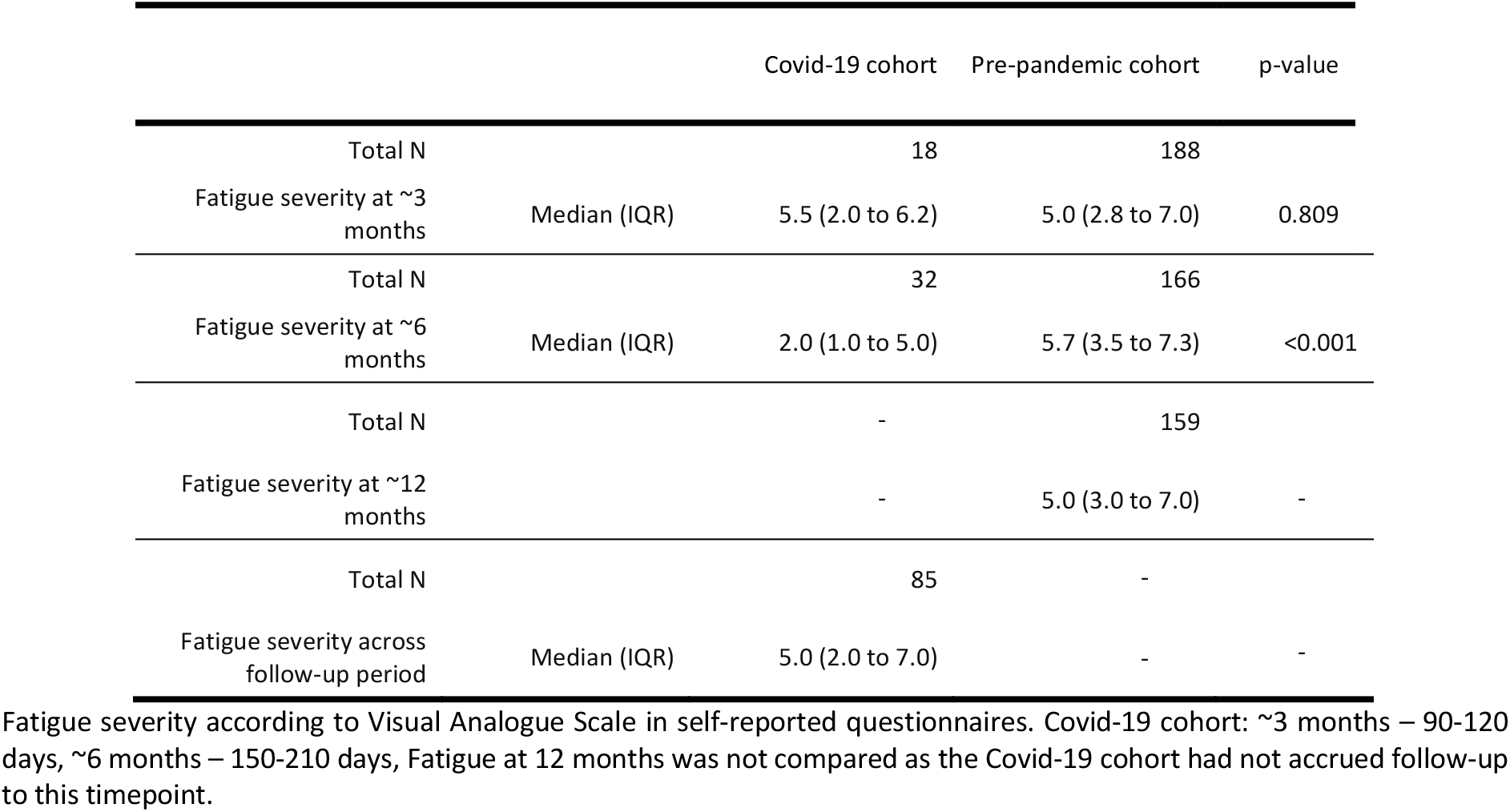
Fatigue severity in Covid-19 and pre-pandemic IMV-patients.

Severe fatigue at the timepoint they responded was reported in 28.2% [24/85] of Covid-19 IMV-patients who responded (Supplementary Table-1). At 3-months post-hospital discharge, the prevalence of severe fatigue was similar in Covid-19 (38.9%, [7/18]) and pre-pandemic IMV-patients (27.1%, [51/188]), and at 6-months significantly less Covid-19 IMV-patients experienced severe fatigue (10.3% [3/29] vs 32.5% [54/166], p=0.015).

### Secondary outcomes

The median MRC grade reported by Covid-19 IMV-patients was 2/5, IQR:1 to 3 (Table 4). At 6-months, the median breathlessness VAS score reported by pre-pandemic IMV-patients was 3.6/10, IQR:1.2 to 6.0 (Supplementary Table-2). The median overall health state reported by Covid-19 IMV-patients was 0.8/1, IQR:0.7 to 0.9. At 6-months, the median MCS and PCS reported by pre-pandemic IMV-patients was 42.5, IQR:34.3 to 54.2, and 35.9, IQR:25.9 to 43.5.

### Predictors of fatigue

At a univariable level, in the Covid-19 group, females under 50 reported more severe fatigue (mean 5.20, 95%CI: 2.93 to 7.47); In pre-pandemic IMV-patients, sex did not affect outcomes (Supplementary Figure-1, Supplementary Table-6).

In Covid-19 IMV-patients, females younger than 50 were nearly 3 times more likely to report greater fatigue (adjusted mean difference 2.56, 95%CI:-0.25 to 5.36, p=0.037). Having prior comorbidity was associated with worse fatigue and QOL (Table-3a).

**Table-3:**
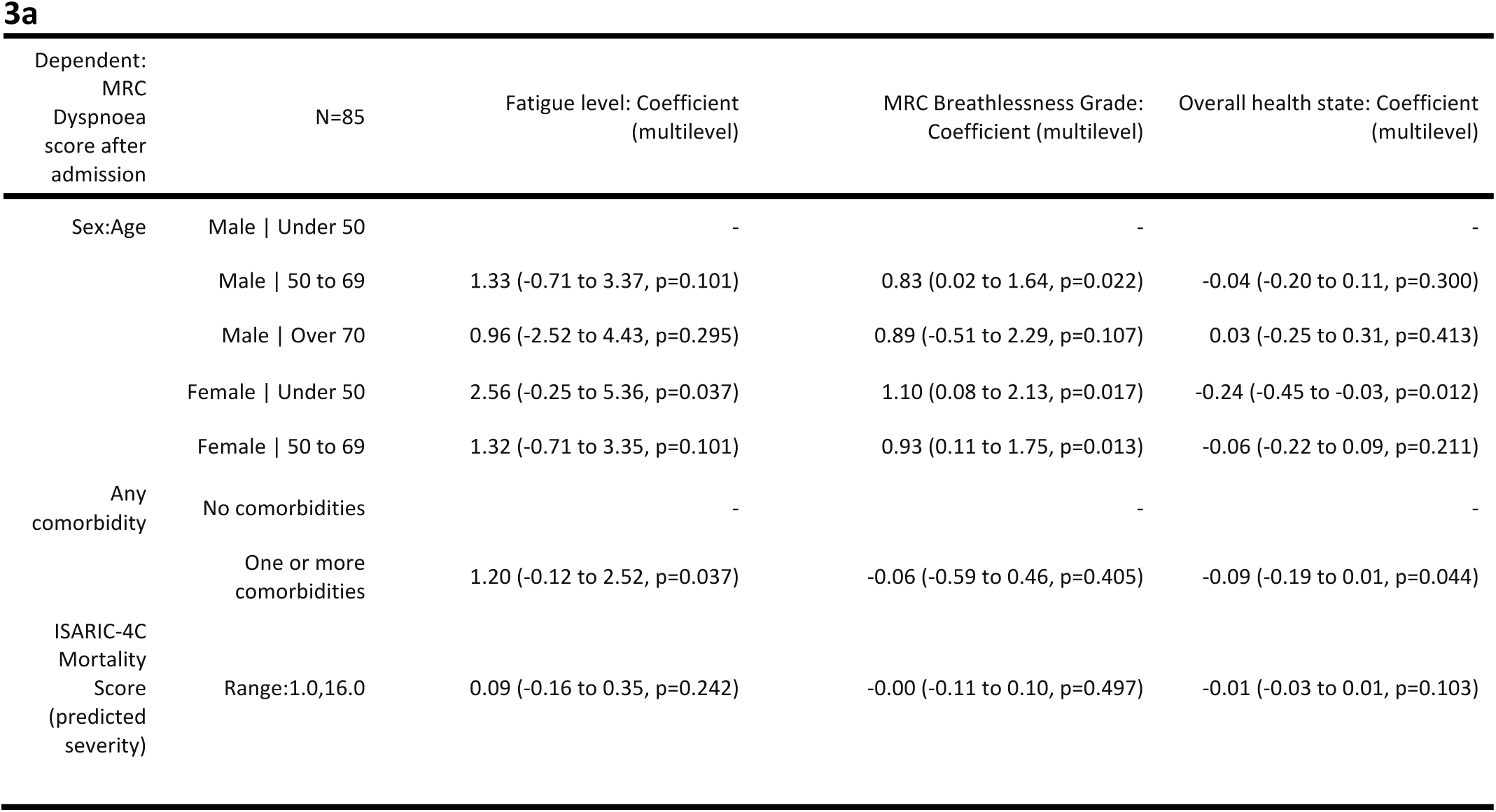
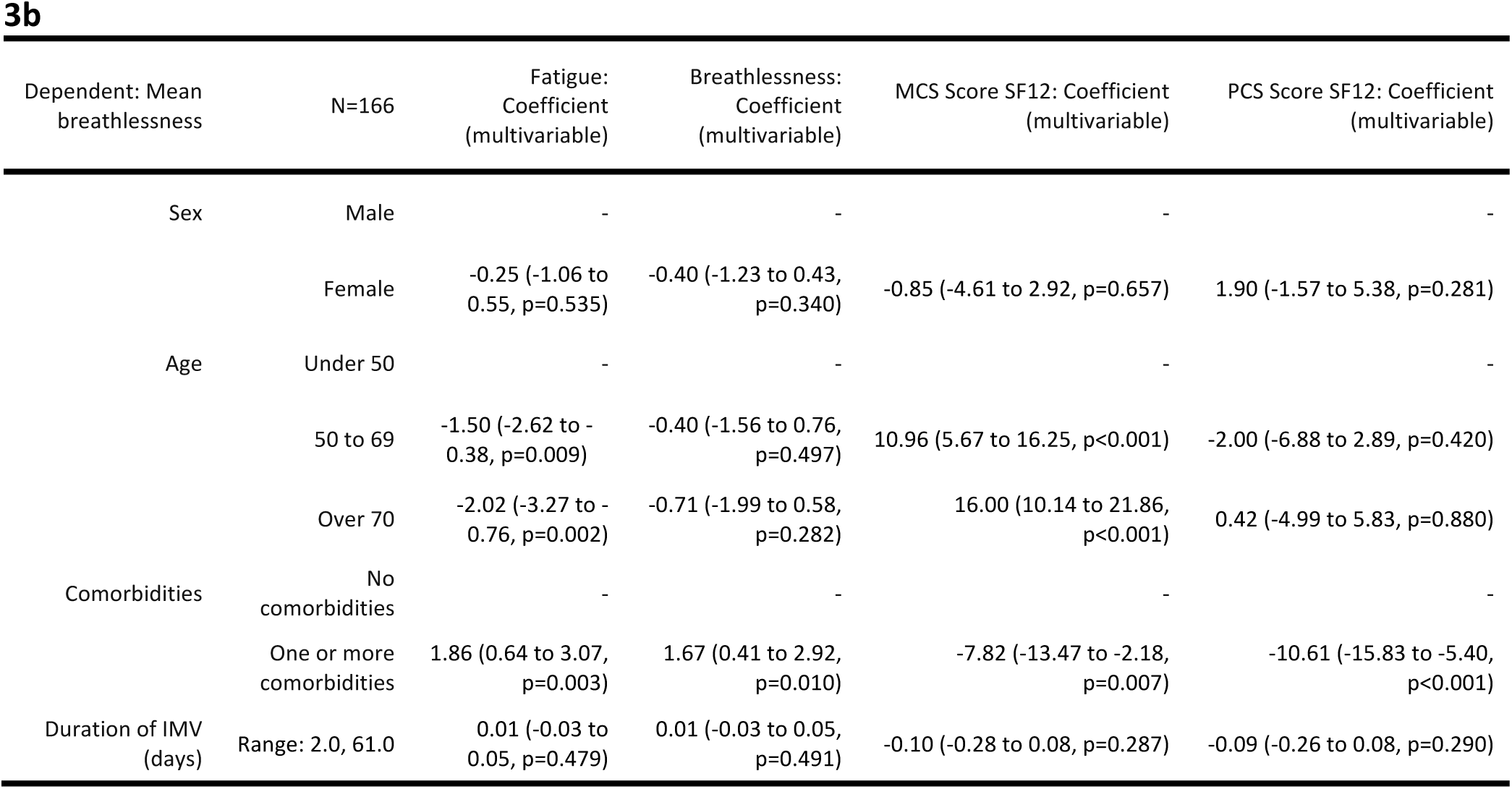
Multilevel regression models for fatigue level, breathlessness level, and quality of life. 3a – Covid-19 cohort, median response (185 days, IQR: 136.8 to 241.2). 3b – Pre-pandemic cohort, at 6-months post-hospital discharge. 3a Model metrics: For Fatigue: Number in model = 85, Number of groups = 27, Log likelihood = −198.73, REML criterion = 397.5. For Breathlessness: Number in model = 88, Number of groups = 27, Log likelihood = − 136.18, REML criterion = 272.4 For Overall health state: Number in model = 91, Number of groups = 27, Log likelihood = − 5.12, REML criterion = 10.2 3b. For Breathlessness: Number in model = 167, Log-likelihood = −397.61, AIC = 809.2, R-squared = 0.049, Adjusted R-squared = 0.019. For Fatigue: Number in model = 166, Log-likelihood = −389.8, AIC = 793.6, R-squared = 0.099, Adjusted R-squared = 0.071 For MCS: Number in model = 165. Log-likelihood = −640.61, AIC = 1295.2, R-squared = 0.18, Adjusted R-squared = 0.15 For PCS: Number in model = 165, Log-likelihood = −627.41, AIC = 1268.8, R-squared = 0.11, Adjusted R-squared = 0.078

In pre-pandemic IMV-patients, sex and duration of IMV showed no significant effect on any outcomes included at 3-12 months (Table-3b, Supplementary Table-3). The presence of comorbidity was significantly associated with worse fatigue at 3-6 months; 3-months adjusted mean difference 1.74, 95%CI:0.63 to 2.85 (p=0.002), 6-months adjusted mean difference 1.86, 95%CI:0.64 to 3.07. Being over 70 was associated with significantly less fatigue at 3 (adjusted mean difference −1.74, 95%CI:-2.86 to −0.61, p=0.003) and 6-months (adjusted mean difference −2.02, 95%CI:-3.27 to −0.76, p=0.002).

In the total sample included (Covid-19 and pre-pandemic patients), having Covid-19 was significantly associated with less severe fatigue (cut-off 7/10, adjusted OR 0.35 (95%CI:0.15 to 0.76, p=0.01) (Figure-2, Supplementary Table-4). Ordinal logistic regression analysis showed that ventilated patients with Covid-19 (adjusted OR 0.50, 95%CI:0.29to 0.84) had less severe fatigue than patients ventilated due to other aetiologies. One or more comorbidities (adjusted OR 3.17, 95%CI:1.75 to 5.81), and age over 70 (adjusted OR 0.39, 95%CI:0.18 to 0.84) were associated with greater fatigue (Supplementary Table-5).

**Figure 2:**
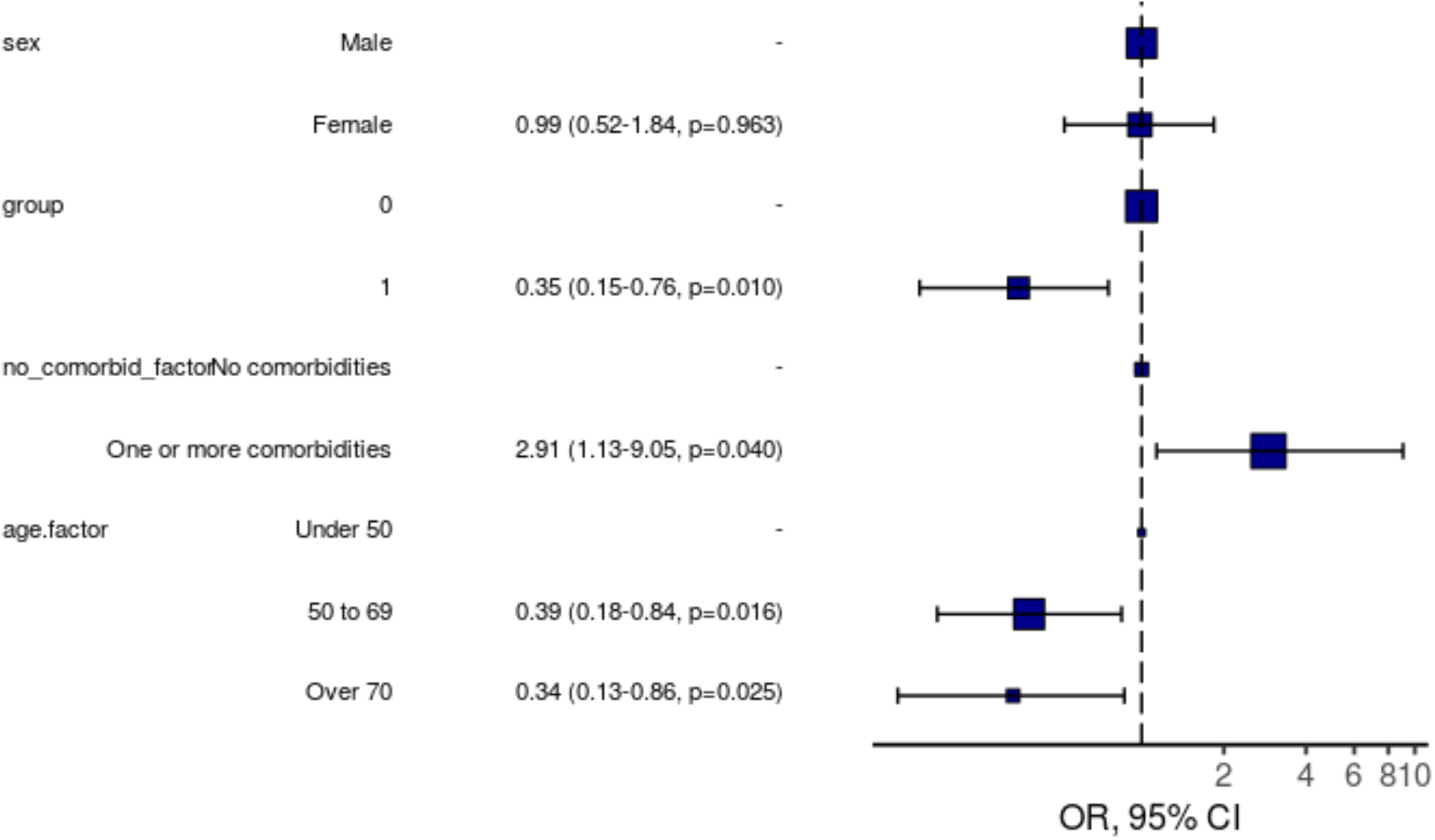
Logistic regression model for severe (>=7/10) fatigue at 6-months including all patients (Covid-19 and pre-pandemic group). OR, 95% CI, p-value. Pre-pandemic patients included at 6-months follow-up. Number in model = 251, AIC = 262.4, C-statistic = 0.676, H&L = Chi-sq(8) 2.26 (p=0.972)

## Discussion

We found a high prevalence and severity of patient-reported persistent fatigue following hospital discharge in ventilated patients with both Covid-19 and non-Covid-19 critical illness. At 3-months post-hospital discharge, the prevalence and severity of fatigue reported was similar following Covid-19 and non-Covid-19 critical illness, however by 6-months, patients ventilated for Covid-19 had significantly less severe fatigue than patients ventilated for other critical illness. In the Covid-19 population, women under 50 experienced more severe fatigue, breathlessness, and worse overall health state compared to other Covid-19 IMV-patients. This contrasted with pre-pandemic IMV-patients where there were no significant sex differences in long-term outcomes. Both Covid-19 and non-Covid-19 critical illness survivors experienced high levels of fatigue, breathlessness, and poor QOL, adding to existing evidence that finding the best rehabilitation services to support ICU-survivors is a research priority^4,5^. In our study, the difference in fatigue prevalence and severity at 6-months, and the difference in factors associated with worse fatigue (female sex), suggest that the recovery trajectory from Covid-19 critical illness may be different to general PICS. Covid-19 critical illness survivors are also more likely to suffer additional respiratory sequelae^31^. In our analysis patients who survived Covid-19 were younger with less prior comorbidity than those surviving other critical illness - this may explain why less fatigue at 6-months was reported after Covid-19 critical illness, as coping with the challenges of comorbidity has been identified as a barrier to ICU-recovery^29,30^. The lower fatigue in Covid-19 patients at 6-months is surprising as Covid-19 patients may have been expected to suffer worse fatigue than pre-pandemic patients if they were more likely to be discharged at a lower functional level due to greater high-demand for ICU beds throughout the pandemic and a focus on survival as opposed to rehabilitation. A multitude of different factors interact to drive fatigue in critical illness survivors, including deconditioning secondary to extended immobilisation, anaemia, poor sleep, depression, post-traumatic stress, respiratory illness (which may be due to prolonged ventilation), chronic disease, and drugs^1,18,29,32–35^. It is possible that once acute illness severity exceeds the threshold for requiring IMV and ICU-admission, further illness severity and underlying disease have little impact on persistent fatigue. This is consistent with recent research which found that post-ICU rehabilitation requirements were unaffected by Covid-19 infection status^36^.

Our study adds to existing literature by highlighting sex differences in recovery from critical illness that are unique to Covid-19. Our finding that women under 50 who survived hospitalisation with acute-Covid-19 experienced worse symptoms and QOL is consistent with other recent evidence in hospitalised cohorts^9,11,12.^ Our study corroborates this in IMV-patients, as previous studies in China and Russia captured small proportions of patients requiring critical care (1%^16^ and 2.6%^12^ respectively).

It is unclear why women under 50 experienced worse fatigue after Covid-19 critical illness. Men may have been more likely to underreport symptoms^37^, but this was not seen in our pre-pandemic control. Women may have been more likely to survive severe acute-Covid-19 disease and therefore to live with worse long-term sequelae^6^, however in our data men survived more severe acute disease. Younger women may have had higher initial exposure to Covid-19, where women are more likely to work in occupations with greater exposure^38^. There are likely to be sex differences in the immune system-response to Covid-19 as susceptibility to severe disease is a consistent feature in many studies ^39^. More research is indicated to elucidate why younger women experience worse long-term outcomes, and if they should be prioritised in vaccination programmes, particularly as the hospitality, education, and healthcare sectors, where females are likely to have greater Covid-19 exposure, are beginning to reopen.

Our data provides important insights into these complex syndromes, which need far greater research to assess the true impact they have on patients. To our knowledge, no previous study has included an appropriate control group with similar illness severity to a Covid-19 cohort. There are several important limitations to our study that must be considered. Firstly, our findings may be subject to responder and survivor bias. The sample sizes included by both CCP-UK and RECOVER were relatively small, and individuals with milder symptoms may have felt less compelled to respond, and those with the most severe symptoms or who died would have been unable to respond. We controlled for CCP-UK and RECOVER measuring outcomes at different timepoints by stratifying fatigue by time since hospital discharge, which allowed equivalent timepoints to be compared but further restricted sample sizes.

Secondly, our results may not be fully representative of all survivors of Covid-19 or non-Covid critical illness, as only patients admitted to a few UK hospitals were included (31 hospitals in CCP-UK, 2 in RECOVER). Thirdly, as CCP-UK and RECOVER did not use the same outcome measures, we were unable to explore differences in acute illness severity (including duration of IMV), breathlessness level, and QOL between Covid-19 and pre-pandemic patients. An ideal study design would have used retrospective measurements of pre-critical illness functional levels, utilised repeated measures (much like the RECOVER study) of those with Covid-19, and would also feature a non-Covid-19 contemporaneous control group, to control for other factors which may impact post-ICU recovery, such as the ‘lockdown’ restrictions in place during CCP-UK’s study period or the level of follow-up care available; Restrictions such as office closures may have meant survivors’ lives were less physically demanding, allowing for greater rest, and the pandemic has heralded the formalisation of ICU-recovery clinics in some areas^40^.

We found high levels of fatigue, breathlessness, and poor QOL following both Covid-19 and non-Covid-19 critical illness. Survivors of Covid-19 critical illness experienced less severe fatigue at 6-months post-hospital discharge than survivors of non-Covid-19 critical illness, potentially due in part to the comparatively younger and less comorbid Covid-19 ICU cohort versus the non-Covid-19 cohort. Younger female survivors experienced more severe fatigue, breathlessness, and worse overall health state compared to other Covid-19 IMV-patients, and future studies should investigate possible mechanisms to explain this. Research targeting interventions to best support critical illness survivors is required in order to optimise recovery.

## Data Availability

This work uses data provided by patients and collected by the NHS as part of their care and support #DataSavesLives. The CO-CIN data was collated by ISARIC4C Investigators. ISARIC4C welcomes applications for data and material access through our Independent Data and Material Access Committee (https://isaric4c.net).

## Funding Statement

This work is supported by grants from: the National Institute for Health Research (NIHR) [award CO-CIN-01], the Medical Research Council [grant MC_PC_19059] and by the NIHR Health Protection Research Unit (HPRU) in Emerging and Zoonotic Infections at University of Liverpool in partnership with Public Health England (PHE), in collaboration with Liverpool School of Tropical Medicine and the University of Oxford [award 200907], NIHR HPRU in Respiratory Infections at Imperial College London with PHE [award 200927], NIHR Biomedical Research Centre at Imperial College London [IS-BRC-1215-20013], and NIHR Clinical Research Network for providing infrastructure support for this research. The views expressed are those of the authors and not necessarily those of the NIHR, MRC or PHE.

Study registration ISRCTN66726260.

The ISARIC WHO CCP-UK study was registered at https://www.isrctn.com/ISRCTN66726260 and designated an Urgent Public Health Research Study by NIHR.

### Ethical considerations

Ethical approval was given by the South Central - Oxford C Research Ethics Committee in England (Ref 13/SC/0149), the Scotland A Research Ethics Committee (Ref 20/SS/0028), and the WHO Ethics Review Committee (RPC571 and RPC572, 25 April 2013.

