## Supplementary material for "Recovery from Covid-19 critical illness: a secondary analysis of the ISARIC4C CCP-UK cohort study and the RECOVER trial"

### Supplementary table-1: Severe fatigue in Covid-19 (n=85) and pre-pandemic (At 3-months: n=188, at 6-months: n=166, at 12-months, n=159) IMV-patients.

Numbers are presented as N (%), unless otherwise denoted as a continuous variable. Patients missing did not respond, or for the pre-pandemic cohort did not respond or did not survive to follow-up. The Covid-19 cohort was stratified according to approximate time since hospital discharge at which they responded: ~3 months was defined as 90-120 days, ~6 months was defined as 150-210 days.

| | | | Covid-19 cohort | Pre-pandemic cohort | p-value |
|---|---|---|---|---|---|
| Fatigue >=7/10 on VAS | ~3 months | Yes | 7 (38.9) | 51 (27.1) | 0.285 |
| | | No | 11 (61.1) | 137 (72.9) | |
| | ~6 months | Yes | 3 (10.3) | 54 (32.5) | 0.015 |
| | | No | 26 (89.7) | 112 (67.5) | |
| | ~12 months | Yes | - | 51 (32.1) | - |
| | | No | - | 108 (67.9) | |
| | Across follow-up period | Yes | 24 (28.2) | - | - |
| | | No | 61 (71.8) | - | |

### Supplementary Table-2: Breathlessness and quality of life in Covid-19 (n=85) and pre-pandemic IMV-patients (n=197).

IQR – Interquartile range, presented as 25th to 75th centiles. Numbers are presented as N (%), unless otherwise denoted as a continuous variable. Patients missing did not respond, or for the pre-pandemic cohort did not respond or did not survive to follow-up.

| | | Covid-19 cohort | Pre-pandemic cohort | N |
|---|---|---|---|---|
| MRC Dyspnoea Grade | Median (IQR) | 2.0 (1.0 to 3.0) | - | 85 |
| Overall health state (ED5D-5L) | Median (IQR) | 0.8 (0.7 to 0.9) | - | 85 |
| Breathlessness at 3 months | Median (IQR) | - | 2.9 (1.1 to 5.0) | 189 |
| Breathlessness at 6 months | Median (IQR) | - | 3.6 (1.2 to 6.0) | 167 |
| Breathlessness at 12 months | Median (IQR) | - | 3.3 (1.1 to 6.1) | 159 |
| MCS (SF-12) at 3 months | Median (IQR) | - | 45.6 (33.3 to 54.7) | 197 |
| MCS (SF-12) at 6 months | Median (IQR) | - | 42.5 (34.3 to 54.2) | 165 |
| MCS (SF-12) at 12 months | Median (IQR) | - | 44.5 (35.1 to 54.3) | 155 |
| PCS (SF-12) at 3 months | Median (IQR) | - | 34.6 (26.7 to 42.9) | 197 |
| PCS (SF-12) at 6 months | Median (IQR) | - | 35.9 (25.9 to 45.3) | 165 |
| PCS (SF-12) at 12 months | Median (IQR) | - | 36.6 (27.5 to 46.2) | 155 |

### Supplementary Table-3: Combined model tables: pre-pandemic IMV-patients.

#### A. 3-months

For breathlessness - Number in model = 189. Log-likelihood = -430.7, AIC = 875.4, R-squared = 0.14, Adjusted R-squared = 0.11. For Fatigue - Number in model = 188, Log-likelihood = -438.43, AIC = 890.9, R-squared = 0.088, Adjusted R-squared = 0.063. For MCS - Number in model = 197, Log-likelihood = -773.64, AIC = 1561.3, R-squared = 0.14, Adjusted R-squared = 0.12. For PCS - Number in model = 197, Log-likelihood = -751.76, AIC = 1517.5, R-squared = 0.037, Adjusted R-squared = 0.012

| Dependent: Mean breathlessness | N=188 | Fatigue: Coefficient (multivariable) | Breathlessness Coefficient (multivariable) | MCS Score SF12: Coefficient (multivariable) | PCS Score SF12: Coefficient (multivariable) |
|---|---|---|---|---|---|
| Sex | Male | - | - | - | - |
| | Female | -0.12 (-0.88 to 0.63, p=0.747) | -0.60 (-1.32 to 0.11, p=0.097) | -0.53 (-4.15 to 3.09, p=0.774) | 1.95 (-1.29 to 5.19, p=0.236) |
| Age | Under 50 | - | - | - | - |
| | 50 to 69 | -1.30 9-2.29 to -0.30, p=0.011) | -0.82 (-1.77 to 0.12, p=0.087) | 7.84 (3.17 to 12.51, p=0.001) | -0.98 (-5.75 to 3.79, p=0.868) |
| | Over 70 | -1.74 (-2.86 to -0.61, p=0.003) | 1.33 (-2.39 to -0.27, p=0.015) | 14.36 (9.03 to 19.69, p<0.001) | -0.98 (-5.75 to 3.79, p=0.868) |
| Comorbidities | No comorbidities | - | - | - | - |
| | One or more comorbidities | 1.74 (0.63 to 2.85, p=0.002) | 2.43 (1.38 to 3.49, p<0.001) | -5.91 (-11.25 to -0.57, p=0.030) | -5.10 (-9.88 to -0.31, p=0.037) |
| Duration of IMV (days) | Range: 2.0, 61.0 | -0.00 (-0.04 to 0.04, p=0.998) | 0.04 (0.00 to 0.08, p=0.028) | 0.10 (-0.07 to 0.28, p=0.257) | -0.05 (-0.21 to 0.11, p=0.530) |

#### B. 12-months

For Breathlessness - Number in model = 159, Log-likelihood = -390.24, AIC = 794.5, R-squared = 0.051, Adjusted R-squared = 0.021 For fatigue - Number in model = 159, Log-likelihood = -380.63, AIC = 775.3, R-squared = 0.033, Adjusted R-squared = 0.0013 For MCS - Number in model = 155, Log-likelihood = -591.98, AIC = 1198, R-squared = 0.13, Adjusted R-squared = 0.098 For PCS - Number in model = 155, Log-likelihood = -606.67, AIC = 1227.3, R-squared = 0.056, Adjusted R-squared = 0.025

| Dependent: Mean breathlessness | N=159 | Fatigue Coefficient (multivariable) | Breathlessness: Coefficient (multivariable) | MCS Score SF12: Coefficient (multivariable) | PCS Score SF12: Coefficient (multivariable) |
|---|---|---|---|---|---|
| Sex | Male | - | - | - | - |
| | Female | 0.04 (-0.83 to 0.91, p=0.920) | 0.08 (-0.80 to 0.97, p=0.853) | -0.96 (-4.62 to 2.69, p=0.604) | 0.06 (-3.95 to 4.08, p=0.975) |
| Age | Under 50 | - | - | - | - |
| | 50 to 69 | 0.47 (-0.86 to 1.80, p=0.489) | 1.11 (-0.23 to 2.46, p=0.103) | 5.80 (0.24 to 11.37, p=0.041) | -4.23 (-10.35 to 1.89, p=0.174) |
| | Over 70 | 0.01 (-1.43 to 1.46, p=0.984) | 0.41 (-1.05 to 1.88, p=0.578) | 12.17 (6.14 to 18.20, p<0.001) | -3.33 (-9.95 to 3.30, p=0.323) |
| Comorbidities | No comorbidities | - | - | - | - |
| | One or more comorbidities | 1.26 (-0.07 to 2.60, p=0.064) | 1.05 (-0.33 to 2.43, p=0.134) | -7.73 (-13.56 to -1.90, p=0.010) | -8.05 (-14.47 to -1.64, p=0.014) |

| Dependent: Mean breathlessness | N=159 | Fatigue Coefficient (multivariable) | Breathlessness: Coefficient (multivariable) | MCS Score SF12: Coefficient (multivariable) | PCS Score SF12: Coefficient (multivariable) |
|---|---|---|---|---|---|
| Duration of IMV (days) | Range: 2.0, 61.0 | -0.02 (-0.06 to 0.02, p=0.397) | -0.03 (-0.07 to 0.01, p=0.142) | 0.04 (-0.13 to 0.21, p=0.611) | 0.02 (-0.16 to 0.21, p=0.796) |

### Supplementary Table-4: Multilevel logistic regression model including all patients (Covid-19 and pre-pandemic group) for self-reported severe fatigue at follow-up (cut-off >7/10).

Number in model = 251, AIC = 262.4, C-statistic = 0.676, H&L = Chi-sq(8) 2.26 (p=0.972) Pre-pandemic patients included at 6-months follow-up.

| Dependent: Fatigue VAS Score | N=251 | Fatigue score <=7 | Fatigue score >7 | OR (univariable) | OR (multivariable) |
|---|---|---|---|---|---|
| Sex | Male | 119 (78.3) | 32 (21.7) | - | - |
| | Female | 74 (74.7) | 25 (25.3) | 1.22 (0.67-2.20, p=0.516) | 0.99 (0.52-1.84, p=0.963) |
| Covid-19 | No | 118 (71.1) | 48 (28.9) | - | - |
| | Yes | 75 (88.2) | 10 (11.8) | 0.33 (0.15-0.66, p=0.003) | 0.35 (0.15-0.76, p=0.010) |
| Any comorbidities | No comorbidities | 47 (90.4) | 5 (9.6) | - | - |
| | One or more comorbidities | 146 (73.4) | 53 (26.5) | 3.41 (1.40-10.23, p=0.014) | 2.91 (1.13-9.05, p=0.040) |
| Age | Under 50 | 28 (63.3) | 16 (36.4) | - | - |
| | 50 to 69 | 126 (80.8) | 30 (19.2) | 0.42 (0.20-0.88, p=0.019) | 0.39 (0.18-0.84, p=0.016) |
| | Over 70 | 39 (76.5) | 12 (23.5) | 0.54 (0.22-1.31, p=0.174) | 0.34 (0.13-0.86, p=0.025) |

### Supplementary Table-5: Ordinal logistic regression model

Residual Deviance: 743.2782; AIC: 761.2782; Number in model = 251

| | Value | Standard error | T value | P value | OR | 95% CI |
|---|---|---|---|---|---|---|
| Covid-19: Yes | -0.69 | 0.27 | -2.59 | 0.477 | 0.50 | 0.29-0.84 |
| Sex: Female | -0.01 | 0.24 | -0.02 | 1.000 | 0.99 | 0.62-1.59 |
| Any comorbidities: One or more | 1.15 | 0.31 | 3.78 | 0.029 | 3.17 | 1.75-5.81 |
| Age: 50-69 | -0.58 | 0.33 | -1.78 | 1.000 | 0.56 | 0.29-1.06 |
| Age: Over 70 | -0.94 | 0.39 | -2.40 | 0.222 | 0.39 | 0.18-0.84 |
| Fatigue level | | | | | | |
| 0-2 \| 2-4 | -1.20 | 0.42 | -2.87 | | | |
| 2-4 \| 4-6 | -0.36 | 0.42 | -0.86 | | | |
| 4-6 \| 6-8 | 0.68 | 0.42 | 1.64 | | | |
| 6-8 \| 8-10 | 2.61 | 0.45 | 5.86 | | | |

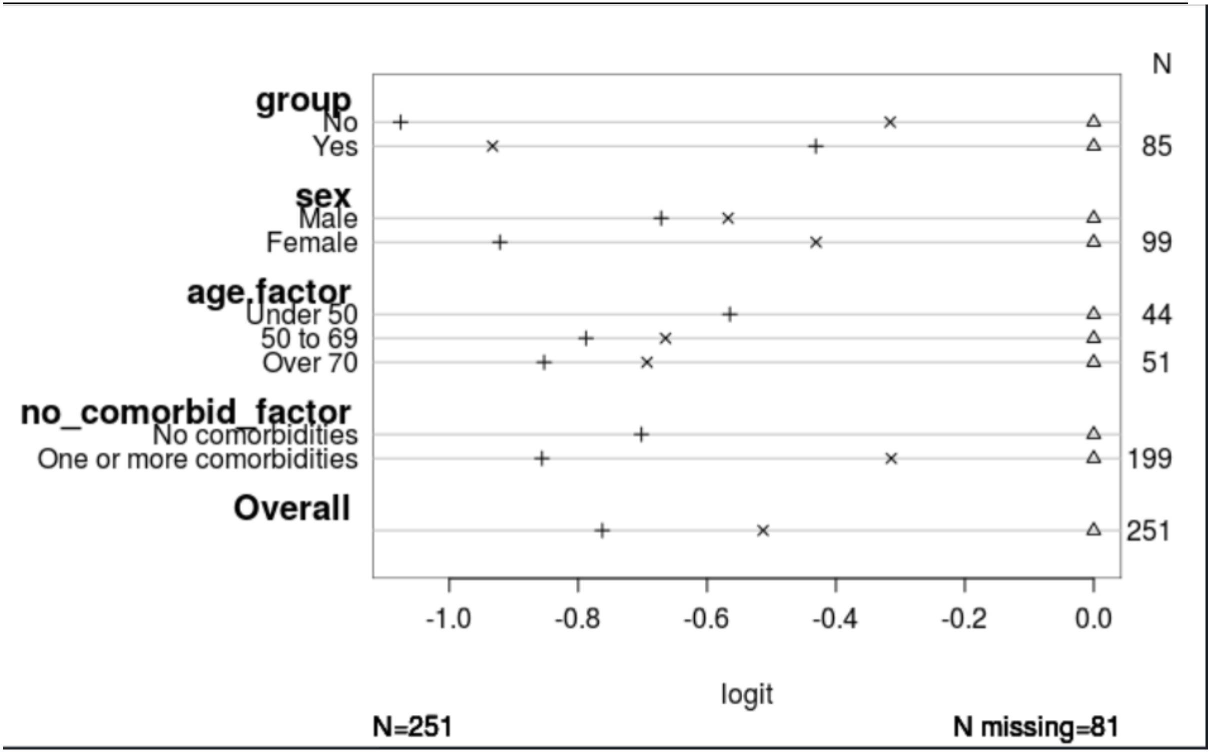


Group – Covid-19 status. Age.factor – age. No_comorbid_factor – Presence of comorbidity.

Supplementary Table-6: Outcomes stratified by age and sex. 6A: Covid-19 cohort. 6B: Pre-pandemic cohort at 3-months post-hospital discharge. 6C: Pre-pandemic cohort at 6-months post-hospital discharge. 6D: Pre-pandemic cohort at 12-months post-hospital discharge.

6A (n=85)

| Age | Sex | Fatigue | Lower limit | Upper limit |
|---|---|---|---|---|
| Under 50 | Male | 2.58 | 0.97 | 4.19 |
| Under 50 | Female | 5.20 | 2.93 | 7.47 |
| 50 to 69 | Male | 4.40 | 3.56 | 5.24 |
| 50 to 69 | Female | 4.40 | 3.22 | 5.58 |
| Over 70 | Male | 4.20 | 1.69 | 6.71 |
| | | MRC Grade | | |
| Under 50 | Male | 1.67 | 1.11 | 2.22 |
| Under 50 | Female | 2.57 | 2.18 | 2.97 |
| 50 to 69 | Male | 2.43 | 2.05 | 2.82 |
| 50 to 69 | Female | 2.70 | 2.25 | 3.15 |
| Over 70 | Male | 2.60 | 1.42 | 3.78 |
| | | Overall Health State | | |
| Under 50 | Male | 0.86 | 0.79 | 0.92 |
| Under 50 | Female | 0.66 | 0.49 | 0.82 |
| 50 to 69 | Male | 0.76 | 0.68 | 0.83 |
| 50 to 69 | Female | 0.72 | 0.59 | 0.84 |
| Over 70 | Male | 0.77 | 0.70 | 0.84 |

6B

Fatigue – N=188. Breathlessness = N=189. MCS – Mental Component Score N=197. PCS – Physical Component Score N=197.

| Age | Sex | Fatigue | Lower limit | Upper limit |
|---|---|---|---|---|
| Under 50 | Male | 5.82 | 4.66 | 6.99 |
| Under 50 | Female | 6.08 | 5.12 | 7.04 |
| 50 to 69 | Male | 4.78 | 4.16 | 5.40 |
| 50 to 69 | Female | 4.59 | 3.59 | 5.60 |
| Over 70 | Male | 4.42 | 3.41 | 5.42 |
| Over 70 | Female | 4.48 | 3.62 | 5.34 |
| | | Breathlessness | | |
| Under 50 | Male | 4.02 | 2.62 | 5.42 |
| Under 50 | Female | 3.80 | 2.31 | 5.29 |
| 50 to 69 | Male | 3.40 | 2.77 | 4.02 |
| 50 to 69 | Female | 2.99 | 2.25 | 3.73 |
| Over 70 | Male | 3.25 | 2.25 | 4.26 |
| Over 70 | Female | 2.60 | 1.87 | 3.32 |
| | | PCS | | |
| Under 50 | Male | 36.72 | 32.23 | 41.20 |
| Under 50 | Female | 37.05 | 31.86 | 42.23 |
| 50 to 69 | Male | 33.42 | 30.63 | 36.21 |
| 50 to 69 | Female | 36.01 | 32.71 | 39.30 |
| Over 70 | Male | 34.84 | 29.84 | 39.84 |
| Over 70 | Female | 35.86 | 32.06 | 39.66 |
| | | MCS | | |
| Under 50 | Male | 36.71 | 30.82 | 42.59 |
| Under 50 | Female | 36.52 | 30.14 | 42.90 |

| | | | | |
|---|---|---|---|---|
| 50 to 69 | Male | 44.75 | 42.14 | 47.36 |
| 50 to 69 | Female | 44.00 | 39.33 | 48.68 |
| Over 70 | Male | 50.66 | 45.97 | 55.35 |
| Over 70 | Female | 49.80 | 44.83 | 54.77 |

## 6C

Fatigue – N=166. Breathlessness = N=167. MCS – Mental Component Score N=165. PCS – Physical Component Score N=165.

| Age | Sex | Fatigue | Lower limit | Upper Limit |
|---|---|---|---|---|
| Under 50 | Male | 6.14 | 4.55 | 7.73 |
| Under 50 | Female | 6.90 | 5.88 | 7.92 |
| 50 to 69 | Male | 5.36 | 4.70 | 6.01 |
| 50 to 69 | Female | 4.82 | 3.94 | 5.70 |
| Over 70 | Male | 4.93 | 3.70 | 6.16 |
| Over 70 | Female | 4.83 | 3.69 | 5.96 |
| | | Breathlessness | | |
| Under 50 | Male | 4.33 | 2.73 | 5.92 |
| Under 50 | Female | 3.65 | 2.14 | 5.17 |
| 50 to 69 | Male | 3.69 | 2.97 | 4.42 |
| 50 to 69 | Female | 3.63 | 2.75 | 4.50 |
| Over 70 | Male | 3.91 | 2.92 | 4.91 |
| Over 70 | Female | 3.22 | 2.19 | 4.26 |
| | | PCS | | |
| Under 50 | Male | 36.45 | 30.19 | 42.71 |
| Under 50 | Female | 39.17 | 33.77 | 44.56 |
| 50 to 69 | Male | 34.61 | 31.40 | 37.82 |
| 50 to 69 | Female | 36.41 | 32.69 | 40.14 |

| | | | | |
|---|---|---|---|---|
| Over 70 | Male | 36.12 | 31.01 | 41.22 |
| Over 70 | Female | 36.69 | 32.57 | 40.81 |
| | | MCS | | |
| Under 50 | Male | 31.75 | 24.82 | 38.69 |
| Under 50 | Female | 33.29 | 27.35 | 39.23 |
| 50 to 69 | Male | 43.83 | 40.66 | 47.01 |
| 50 to 69 | Female | 42.80 | 38.38 | 47.22 |
| Over 70 | Male | 48.62 | 44.25 | 53.00 |
| Over 70 | Female | 45.87 | 40.89 | 50.86 |

## 6D

Fatigue – N=159. Breathlessness = N=159. MCS – Mental Component Score N=155. PCS – Physical Component Score N=155.

| Age | Sex | Fatigue | Lower limit | Upper limit |
|---|---|---|---|---|
| Under 50 | Male | 5.04 | 2.66 | 7.43 |
| Under 50 | Female | 4.53 | 3.24 | 5.82 |
| 50 to 69 | Male | 5.29 | 4.58 | 6.00 |
| 50 to 69 | Female | 5.36 | 4.51 | 6.20 |
| Over 70 | Male | 4.89 | 3.74 | 6.04 |
| Over 70 | Female | 5.13 | 4.04 | 6.23 |
| | | Breathlessness | | |
| Under 50 | Male | 3.52 | 1.33 | 5.72 |
| Under 50 | Female | 2.37 | 0.89 | 3.85 |
| 50 to 69 | Male | 3.85 | 3.07 | 4.62 |
| 50 to 69 | Female | 4.42 | 3.57 | 5.27 |
| Over 70 | Male | 3.68 | 2.61 | 4.74 |
| Over 70 | Female | 3.33 | 2.26 | 4.40 |

| | | | | |
|---|---|---|---|---|
| | | MCS | | |
| Under 50 | Male | 36.65 | 26.01 | 47.28 |
| Under 50 | Female | 39.53 | 33.16 | 45.91 |
| 50 to 69 | Male | 44.56 | 41.52 | 47.59 |
| 50 to 69 | Female | 42.02 | 37.66 | 46.38 |
| Over 70 | Male | 48.81 | 44.95 | 52.66 |
| Over 70 | Female | 49.07 | 45.34 | 52.81 |
| | | PCS | | |
| Under 50 | Male | 37.40 | 29.77 | 45.02 |
| Under 50 | Female | 45.19 | 36.70 | 53.68 |
| 50 to 69 | Male | 37.73 | 34.27 | 41.19 |
| 50 to 69 | Female | 35.44 | 31.52 | 39.36 |
| Over 70 | Male | 36.37 | 30.49 | 42.26 |
| Over 70 | Female | 37.29 | 33.16 | 41.43 |

Supplementary Figure-1: Outcomes stratified by age and sex. Red: female. Blue: male.

1A: Covid-19 cohort (n=85) median response (185 days, IQR: 136.8 to 241.2). A- Fatigue severity, B – Breathlessness, C – Overall health state.

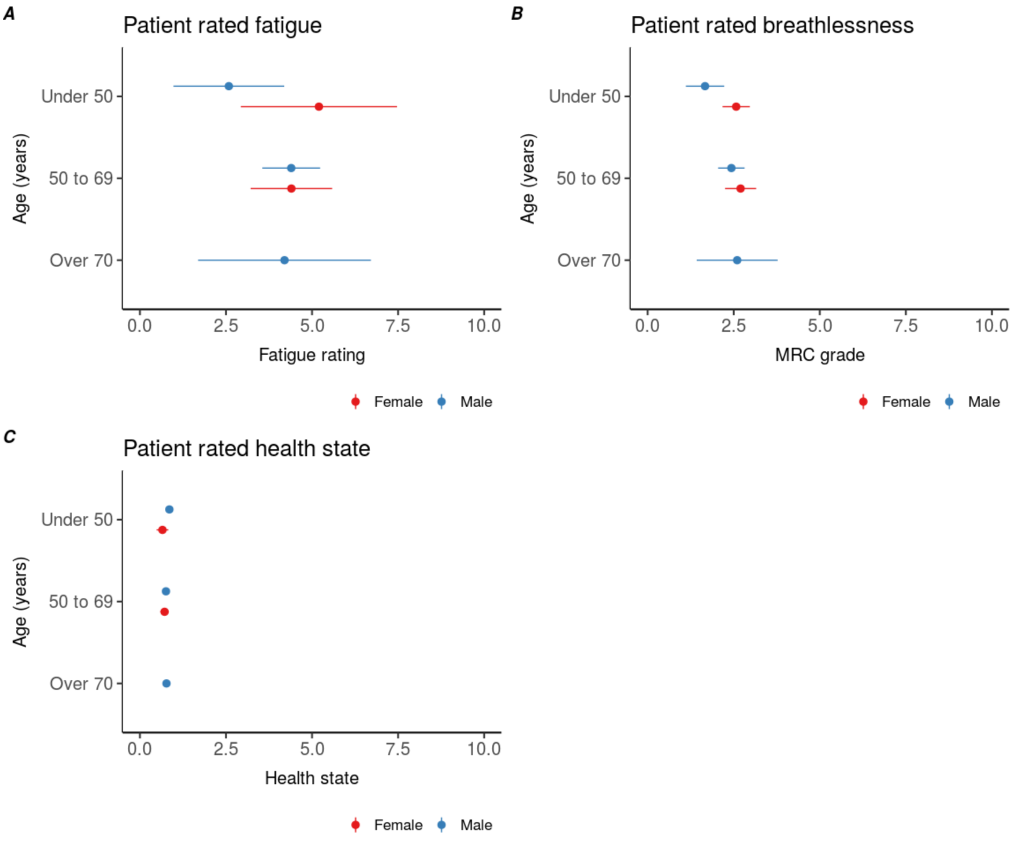

1B-A: Pre-pandemic patients at 3-months post-hospital discharge (n=188). A- Fatigue severity, B – breathlessness, C – Mental Composite Score, D – Physical Composite Score.

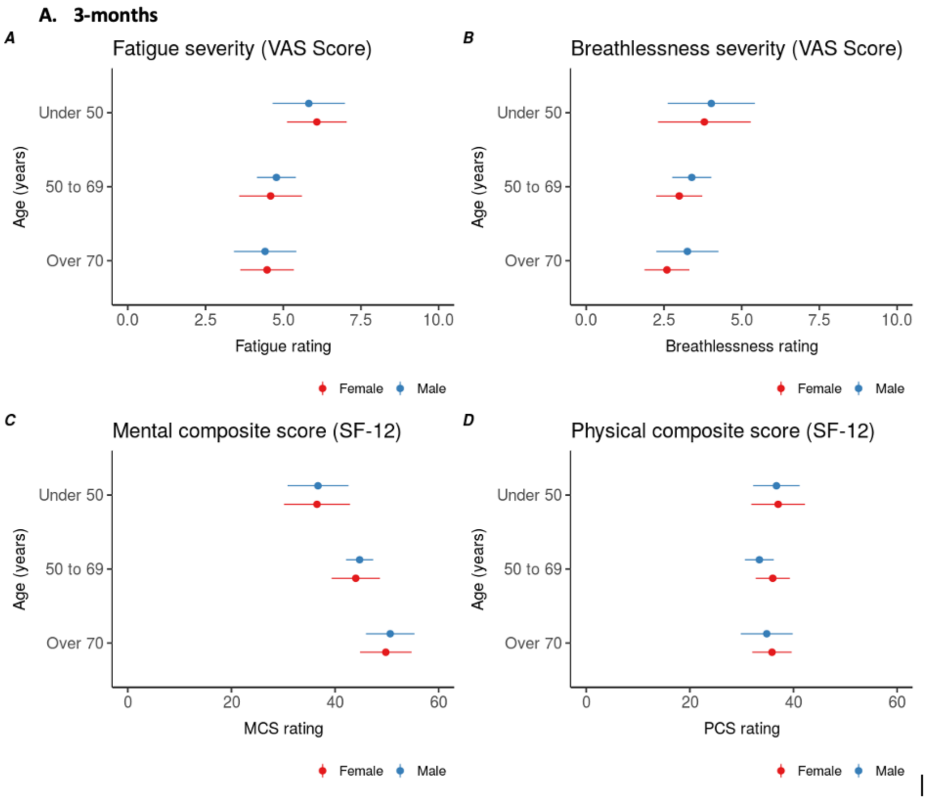


Figure 1B-B: Pre-pandemic patients at 6-months post-hospital discharge (n=166). A- Fatigue severity, B – breathlessness, C – Mental Composite Score, D – Physical Composite Score.

**B. 6-months**

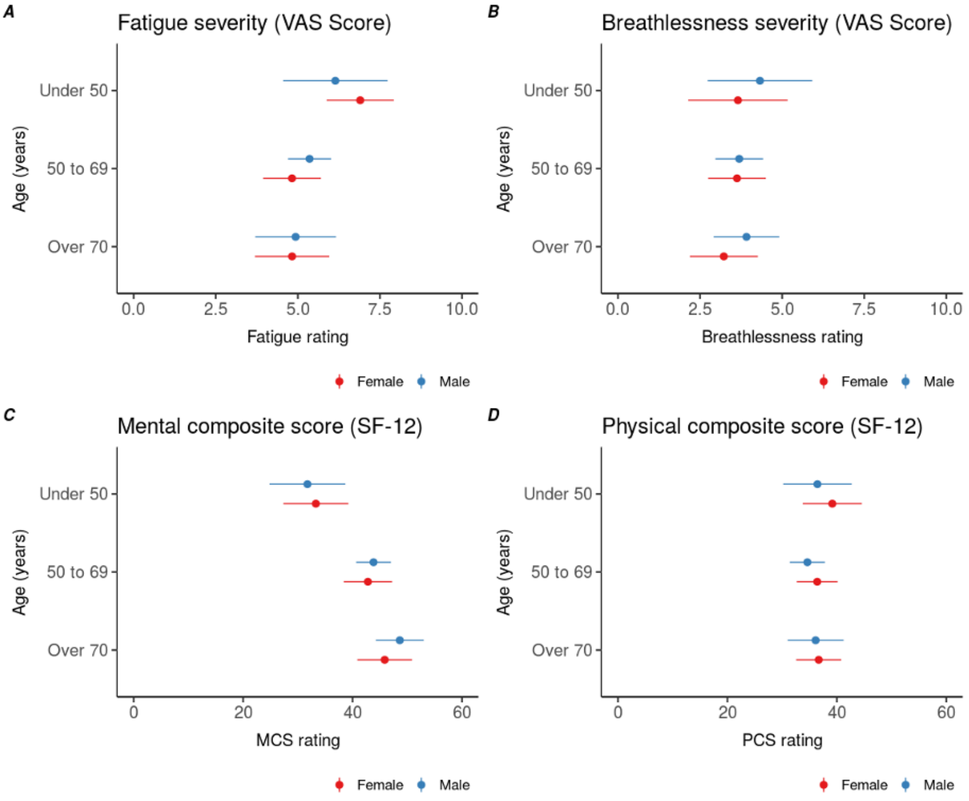


Figure 1B-C: Pre-pandemic patients at 12-months post-hospital discharge (n=159). A- Fatigue severity, B – breathlessness, C – Mental Composite Score, D – Physical Composite Score.

**12-months**

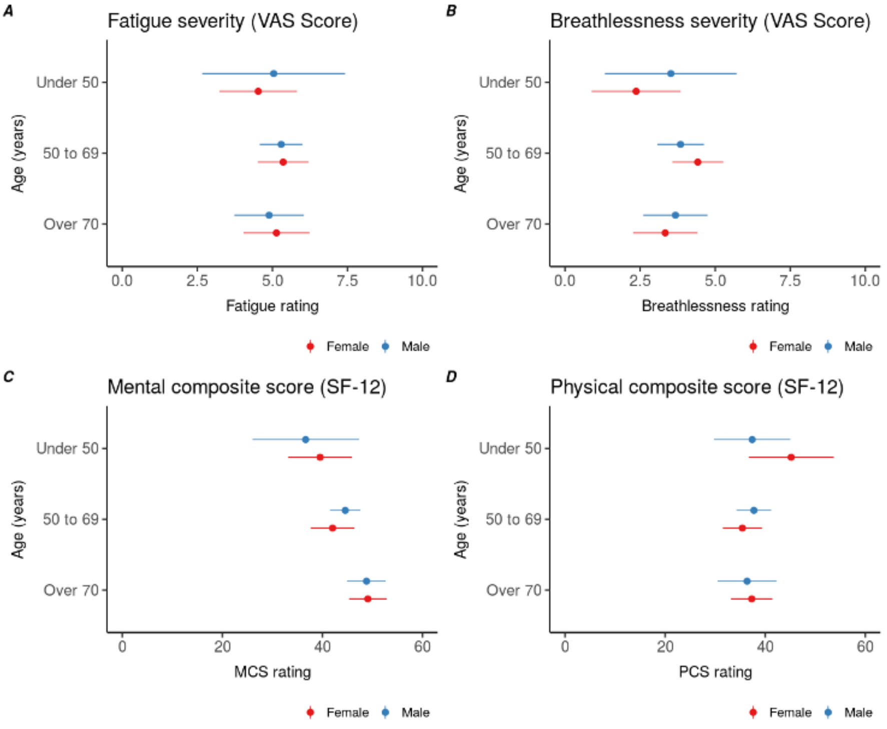
